# A survey investigating United States federal grant submissions proposing to investigate therapeutic applications of psychedelics

**DOI:** 10.1101/2024.10.12.24315367

**Authors:** Brian S. Barnett

## Abstract

This study surveyed researchers to assess the contents and funding success of federal grant applications for research into therapeutic applications of psychedelics in the United States. The author emailed an anonymous survey to the corresponding authors of the 50 most-cited articles on psychedelics published after 2000 and disseminated it via Twitter. Ten researchers responded, reporting on 24 grant submissions for psilocybin, ibogaine, LSD, MDMA, and other psychedelics, all to the National Institutes of Health (NIH), from the early 1990s onward. The number of grant applications rose noticeably starting in 2006. Of all grant applications assessed,16.7% were funded, lower than the NIH’s 23.4% average funding rate for R-01 equivalent grants between 1998-2023. More specifically, while no relevant grant applications submitted prior to 2006-2010 were funded by NIH, the funding rate of applications since then, estimated at 19.05% to 22.2%, is close to the average annual NIH funding rate of 20.6 ± 1.9% for R-01 equivalent grant applications from 2006 to early 2023. Respondents generally believed applications for this line of research had a lower chance of success compared to other lines of research, although they felt the funding landscape has improved in recent years, in line with this study’s other findings.

## Introduction

The therapeutic potential of psychedelics for psychiatric and substance use disorders has generated significant scientific interest in recent decades (Reiff et al. 2020; Singh 2023; Spencer, Miniño, and Warner 2022; Barnett and Weleff 2022). However, since the emergence of the psychedelic renaissance in the early 2000s, financial support for studies on therapeutic applications of psychedelics has largely come from philanthropy and, more recently, the biotechnology industry, rather than governments. For example, from 2006 to 2020 the United States (US) National Institutes of Health (NIH) did not provide direct grant support for a single psychedelic-assisted therapy clinical trial (Barnett, Parker, and Weleff 2021). Notably, in the last few years, governments around the world, including those of Australia, Canada, New Zealand, the United Kingdom, and the US, have begun increasing funding for this area of research.

In October 2021, a multisite clinical trial investigating psilocybin for nicotine use disorder was funded by the National Institute on Drug Abuse (NIDA), an NIH institute, becoming the first contemporary federally funded psychedelic-assisted therapy study in the US (Johns Hopkins Medicine 2021). Since then, NIDA has issued requests for applications (RFAs) for investigations of psychedelics’ therapeutic potential for substance us disorders (National Institutes of Health 2023a) and the National Institute of Mental Health (NIMH), another NIH institute, has issued guidance on grant applications for both animal and human studies of psychedelics’ therapeutic applications (National Institutes of Health 2022a).

Over the nearly half century preceding these developments, federally supported psychedelic studies in the US had primarily focused on studying these compounds as drugs of misuse despite their limited addictive potential (Shalit, Rehm, and Lev-Ran 2019). Prior to this, NIH, the largest public funder of biomedical research in the world (National Institutes of Health 2023b), had extensively funded therapeutic research into lysergic acid diethylamide (LSD) from the 1950s to early 1970s (National Institutes of Health 1955; Oram 2014), when NIMH concluded LSD had no therapeutic applications (Cohen and Krippner 1985). Afterwards, federally funded studies involving administration of psychedelics to human subjects ceased until NIDA funded a pharmacological investigation of dimethyltryptamine (DMT) in the early 1990s (Strassman 1996). NIDA also funded animal studies on the anti-addictive potential of ibogaine in the early 1990s, with favorable results, though the organization refused to fund subsequent proposed human trials (Oaklander 2021).

Given the changing landscape in federal funding for studies of therapeutic applications of psychedelic in the United States, it is important to assess how funding levels for this line of research have changed over time, as well as what kinds of studies are being proposed. Unfortunately, there are no published studies on grant applications for therapeutic psychedelic studies made to US federal agencies. While some information about federally funded grants in the US must be made publicly available via resources such as NIH Reporter and Freedom of Information Act (FOIA) requests, unfunded federal grant applications are considered proprietary and not publicly accessible, even via FOIA request (US National Institutes of Health 2023). Therefore, it is not possible for those outside of relevant federal agencies to even determine if unfunded grant applications for therapeutic psychedelic studies exist, making this topic particularly challenging to study. However, it is still possible to survey researchers who have submitted grant applications to federal agencies, provided one can determine who some of these people might be. Therefore, the purpose of this study was to gather data on grant applications submitted to US federal agencies for therapeutic psychedelic studies via an anonymous survey of authors of high impact psychedelic research articles that was also disseminated via Twitter and snowball sampling.

## Methods

### Survey instrument

The survey was conducted using an anonymous online survey. Regarding terminology, the survey instrument contained the following guidance for respondents, “For the purposes of this survey, therapeutic psychedelic studies include in vitro, animal, human, and other types of studies that focus on applications of psychedelics potentially beneficial to human health. These do not include studies focusing on addictive or other negative aspects of psychedelics except as these issues relate to therapeutic applications of psychedelics. Ketamine and marijuana should not be considered psychedelics for this survey.” Any researcher who had submitted a grant application to a US federal agency for a study of potential therapeutic applications of psychedelics, whether the grant application was funded or unfunded, was eligible to participate. Data from the survey were collected and managed using REDCap electronic data capture tools hosted at Cleveland Clinic (Harris et al. 2009).

The first section of the survey inquired about demographic information and the second section asked respondents about relevant grant applications, with a subsection focusing on submissions to NIH. The third section asked respondents to register their opinions about research funding in this area using a seven-point Likert-scale. The final section posed two questions, one asking how respondents dealt with obtaining funding for projects where grant applications to federal agencies were rejected and the other asking them to provide any additional information about their experiences with attempting to obtain federal funding for therapeutic psychedelic studies.

### Study population and survey dissemination

The survey was emailed to corresponding authors of the 50 most highly cited articles on psychedelics with a U.S. based corresponding author. The author searched for the keyword “psychedelic” using Web of Science on April 1, 2023, and contacted qualifying corresponding authors via email on April 3, 2023. A reminder email was sent two weeks later. On April 4, 2023, the survey was also posted on the author’s Twitter account. On the survey website and in the introductory email, participants were encouraged to participate in snowball sampling by forward the survey onto their contacts. The survey remained open until August 13, 2023.

### Ethics

This study was declared exempt by the Cleveland Clinic Institutional Review Board. Certain measures, such as not reporting data on grant applications for individual psychedelics and reporting years of grant applications in blocks, were undertaken to protect respondent anonymity.

### Statistical analysis

Descriptive statistics were calculated in Microsoft Excel (Redmond, Washington). Some participants left certain questions blank. There was no substitution for missing data. Calculations include only those participants who provided a response to a particular question.

## Results

There were 10 respondents. A response rate could not be calculated since it is unknown how many people receiving a survey invitation or seeing it online had submitted a relevant grant application to a US federal agency and would qualify to take the survey. The typical respondent was a middle aged, White, male, pharmacologist who had received significant grant funding over their career from multiple NIH institutes. Demographic details are listed in Table 1. On average, respondents had submitted two grants for therapeutic psychedelic studies to federal government agencies. Respondents reported submitting relevant applications as far back as the 1991-1995 time block. One respondent did not provide data on the number of relevant grant applications submitted or the federal agencies to which they submitted their grant applications to. However, among the 9 who did, all submitted their applications to NIH, most frequently to NIMH and NIDA. They most frequently proposed to study therapeutic effects of psilocybin, though applications had also been submitted for LSD, 3,4-Methylenedioxymethamphetamine (MDMA), ibogaine, and other psychedelics. For further details on grant applications, see Table 2.

**Table 1.**
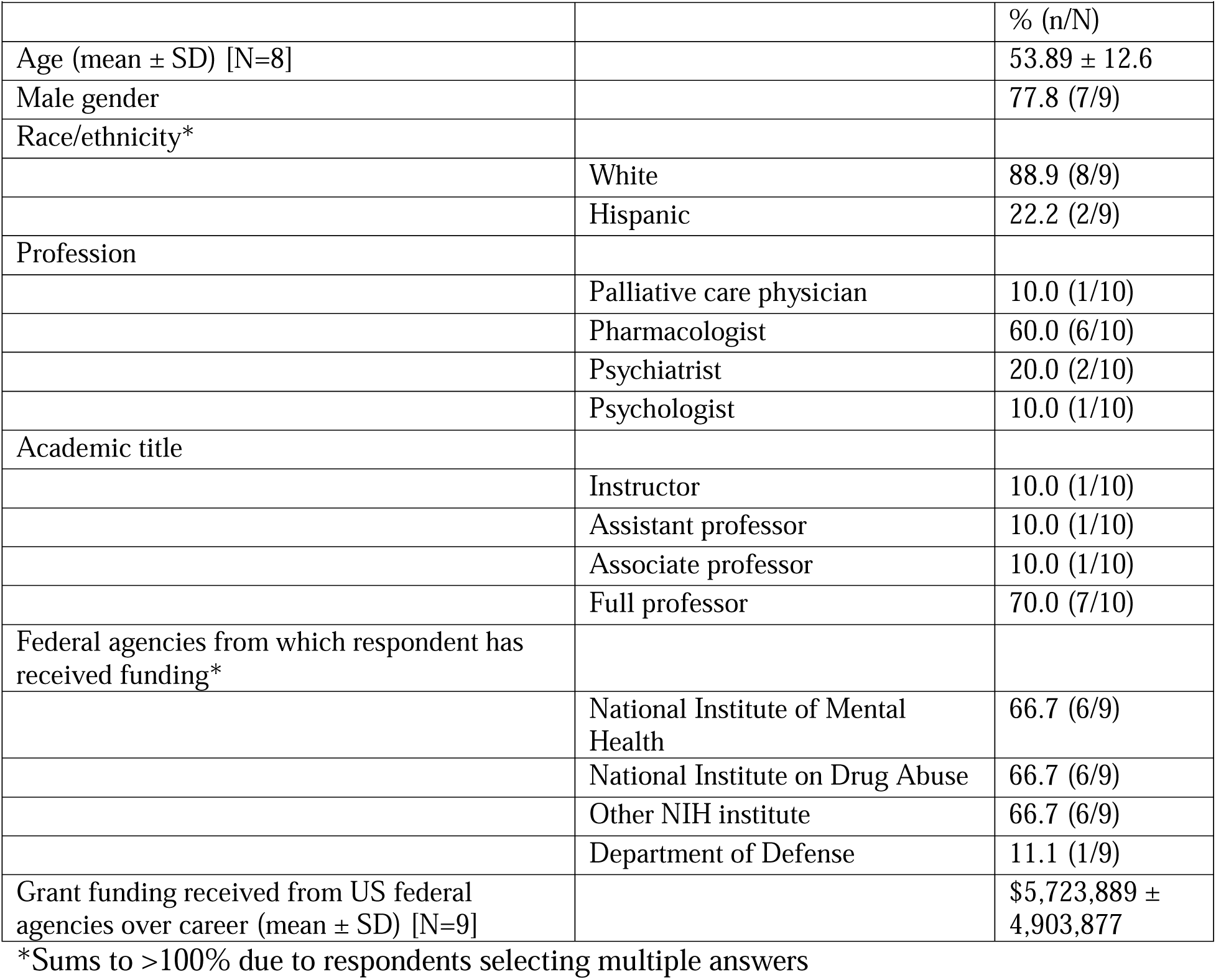
Respondent demographic information.

| **Table 1. Respondent demographic information** |  |  |
| --- | --- | --- |
|  |  | % (n/N) |
| Age (mean ± SD) [N=8] | | 53.89 ± 12.6 |
| Male gender |  | 77.8 (7/9) |
| Race/ethnicity* |  |  |
|  | White | 88.9 (8/9) |
|  | Hispanic | 22.2 (2/9) |
| Profession |  |  |
|  | Palliative care physician | 10.0 (1/10) |
|  | Pharmacologist | 60.0 (6/10) |
|  | Psychiatrist | 20.0 (2/10) |
|  | Psychologist | 10.0 (1/10) |
| Academic title |  |  |
|  | Instructor | 10.0 (1/10) |
|  | Assistant professor | 10.0 (1/10) |
|  | Associate professor | 10.0 (1/10) |
|  | Full professor | 70.0 (7/10) |
| Federal agencies from which respondent has received funding* |  |  |
|  | National Institute of Mental Health | 66.7 (6/9) |
|  | National Institute on Drug Abuse | 66.7 (6/9) |
|  | Other NIH institute | 66.7 (6/9) |
|  | Department of Defense | 11.1 (1/9) |
| Grant funding received from US federal agencies over career (mean ± SD) [N=9] | | \$5,723,889 ± 4,903,877 |

\*Sums to >100% due to respondents selecting multiple answers

**Table 2.**
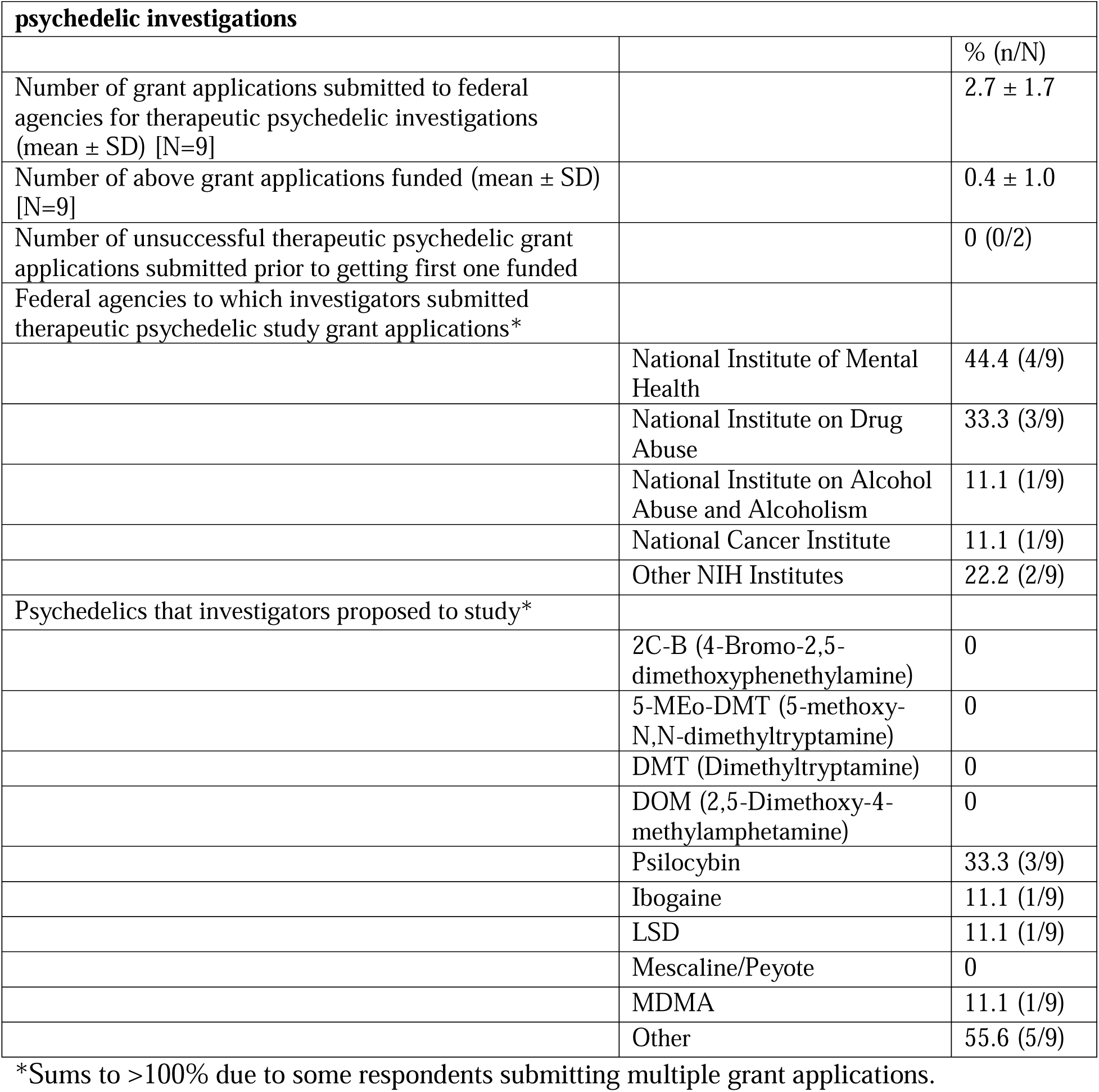
Information about grant applications submitted to federal agencies for therapeutic.

| **psychedelic investigations** |  |  |
| --- | --- | --- |
|  |  | % (n/N) |
| Number of grant applications submitted to federal agencies for therapeutic psychedelic investigations (mean ± SD) [N=9] | | 2.7 ± 1.7 |
| Number of above grant applications funded (mean ± SD) [N=9] | | 0.4 ± 1.0 |
| Number of unsuccessful therapeutic psychedelic grant applications submitted prior to getting first one funded |  | 0 (0/2) |
| Federal agencies to which investigators submitted therapeutic psychedelic study grant applications* |  |  |
|  | National Institute of Mental Health | 44.4 (4/9) |
|  | National Institute on Drug Abuse | 33.3 (3/9) |
|  | National Institute on Alcohol Abuse and Alcoholism | 11.1 (1/9) |
|  | National Cancer Institute | 11.1 (1/9) |
|  | Other NIH Institutes | 22.2 (2/9) |
| Psychedelics that investigators proposed to study* |  |  |
|  | 2C-B (4-Bromo-2,5-dimethoxyphenethylamine) | 0 |
|  | 5-MEO-DMT (5-methoxy-N,N-dimethyltryptamine) | 0 |
|  | DMT (Dimethyltryptamine) | 0 |
|  | DOM (2,5-Dimethoxy-4-methylamphetamine) | 0 |
|  | Psilocybin | 33.3 (3/9) |
|  | Ibogaine | 11.1 (1/9) |
|  | LSD | 11.1 (1/9) |
|  | Mescaline/Peyote | 0 |
|  | MDMA | 11.1 (1/9) |
|  | Other | 55.6 (5/9) |

\*Sums to >100% due to some respondents submitting multiple grant applications.

Studies were primarily in vitro or animal focused, though multiple human investigations, including one psychedelic-assisted therapy trial, had been proposed. The three most common proposed conditions for study were major depressive disorder, alcohol use disorder, and cocaine use disorder. 20% (N=2/10) of respondents reported receiving federal funding for at least one therapeutic psychedelic study and, notably, neither of these respondents reported any unsuccessful therapeutic psychedelic grant applications submissions to federal agencies prior to getting their first therapeutic psychedelic grant application funded from one. For further details on types of studies proposed and diagnoses for which therapeutic applications of psychedelics were to be investigated, see Table 3.

**Table 3.** Data on proposed study type and diagnoses of focus in grant applications for therapeutic psychedelic studies submitted to federal agencies.

|  |  |
| --- | --- |
|  | % (n/N) |
| **Type of study proposed\*** |  |
| In vitro | 70 (7/10) |
| Animal | 70 (7/10) |
| Human- psychedelic-assisted therapy | 10 (1/10) |
| Human- neuroimaging | 0 |
| Human- pharmacology | 10 (1/10) |
| Human- other type of study | 20 (2/10) |
| **Diagnoses proposed\*** |  |
| Alcohol use disorder | 30 (3/10) |
| Cocaine use disorder | 30 (3/10) |
| Major depressive disorder | 40 (4/10) |
| Obsessive compulsive disorder | 10 (1/10) |
| Posttraumatic stress disorder | 10 (1/10) |
| Stimulant use disorder | 20 (2/10) |
| Other condition | 20 (2/10) |

\*Sums to >100% due to some respondents submitting multiple grant applications

Of the 24 grant applications submitted, 16.7% (N=4/24) received federal funding. For years in which grant applications were submitted, see Table 4. Note that since participants were asked in what year they submitted at least one therapeutic psychedelic study grant proposal, the total N is 21 for this table instead of 24. This is relevant to the subsequent discussion about years in which applications were funded and associated funding rates. Though the survey did not specifically ask about years in which grant applications were funded, the two respondents who were successful in achieving grant funding did not submit their first relevant grant applications until the 2006-2010 time block, allowing the deduction that no grant applications were funded from the 1991-1995 time block to the 2001-2005 time blocks, a period in which at least 3 applications but no more than six were submitted. These two successful applicants also submitted grant applications in more recent years, so it is not possible from these data to deduce exactly when from 2006 to early 2023 the first grant applications in the sample were funded. However, if we assume the first funded application was in the 2006-2010 time block, and we assume that 21 of the 24 applications were submitted from 2006 to early 2023, this would yield a funding rate of 19.05% (4/21) during that period. If we assume that 18 applications were submitted during that period, the funding rate would increase to 22.2% (4/18).

**Table 4.** Years in which grant applications proposing to study therapeutic applications of psychedelics were submitted*.

|  |  |
| --- | --- |
| Years | Proportion of applications submitted<br>% (n/N) |
| 2021-early 2023 | 23.8 (5/21) |
| 2016-2020 | 23.8 (5/21) |
| 2011-2015 | 14.3 (3/21) |
| 2006-2010 | 23.8 (5/21) |
| 2001-2005 | 0 |
| 1996-2000 | 9.5 (2/21) |
| 1991-1995 | 4.8 (1/21) |
| 1990 and prior | 0 |

\*Due to some investigators submitting multiple grants in the same time period, N=21 instead of 24.

90.0% (9/10) of respondents reported at least one unfunded grant application for a therapeutic psychedelic study made to a federal agency. 22.2% (2/9) of the participants who submitted relevant grant applications to NIH received funding for at least one application. Among the participants who did not receive NIH funding for at least one grant application, 71.4% (N=5/7) reported receiving an overall impact score. An impact score is a measure of application quality made by a review committee. Applications rejected early in the grant application process are not sent to a review committee and therefore do not receive an impact score (see discussion for more details on score interpretation). The best overall impact score (mean ± standard deviation) of a grant application to NIH that was not funded was 36.3 ± 11.2 [N=4]. The best overall impact score of a grant application to NIH that was funded was 22.5 ± 6.4 (N=2).

Respondents predominantly felt that grant applications for therapeutic studies of psychedelics were significantly less likely to receive federal grant compared to other grant applications they had submitted and that the US federal government is significantly underinvested in therapeutic psychedelic research. Most respondents believed the odds of receiving federal grant funding for therapeutic psychedelic studies have improved over the last five years, with the qualifier “somewhat” being most frequently selected by these individuals. Of the 9 respondents who had at least one unfunded NIH grant application, 33.3% (N=3) reported their proposed study was still unfunded. The rest obtained financial support from alternate sources, with funding from their own institution and psychedelic-focused non-profit organizations being most frequently reported. For further details on answers to these questions, see Table 5.

**Table 5.** Respondent answers to opinion questions.

| **Table 5. Respondent answers to opinion questions** |  |
| --- | --- |
|  | % (n/N) |
| **Have you noticed any difference in how frequently your therapeutic psychedelic grant applications receive US federal agency funding compared to your grant applications for research in other areas?** |  |
| Grant applications for therapeutic psychedelic studies appear: |  |
| Significantly less likely to receive federal grant funding | 88.9 (8/9) |
| Moderately less likely to receive federal grant funding | 0 |
| Somewhat less likely to receive federal grant funding | 11.1 (1/9) |
| To receive federal grant funding at the same rate as other areas of research | 0 |
| Somewhat more likely to receive federal grant funding | 0 |
| Moderately more likely to receive federal grant funding | 0 |
| Significantly more likely to receive federal grant funding | 0 |
| **How do you view the current United States federal government investment in therapeutic psychedelic research given existing data on therapeutic efficacy and safety?** |  |
| Significantly overinvested | 10.0 (1/10) |
| Moderately overinvested | 0 |
| Somewhat overinvested | 0 |
| Appropriately invested | 0 |
| Somewhat underinvested | 0 |
| Moderately underinvested | 10.0 (1/10) |
| Significantly underinvested | 80.0 (8/10) |
| **How do you think the odds of receiving federal grant funding for therapeutic psychedelic research have changed over the last 5 years?** |  |
| Worsened significantly | 0 |
| Worsened moderately | 0 |
| Worsened somewhat | 0 |
| Unchanged | 30.0 (3/10) |
| Improved somewhat | 40.0 (4/10) |
| Improved moderately | 20.0 (2/10) |
| Improved significantly | 10.0 (1/10) |
| **If you have had US federal grant applications for therapeutic psychedelic research rejected, how have you dealt with getting those projects funded (choose all that apply)?\*** |  |
| Project or projects remain unfunded | 33.3 (3/9) |
| Obtained internal funding from own institution | 55.6 (5/9) |
| Obtained funding from crowdsourcing | 0 |
| Obtained funding from psychedelic focused non-profit organizations | 44.4 (4/9) |
| Obtained funding from other types of non-profit organizations | 33.3 (3/9) |
| Self-funded | 11.1 (1/9) |
| Obtained funding from other sources | 55.6 (5/9) |

\*Sums to >100% due to respondents being able to select multiple answers

Multiple respondents provided written responses about their experiences attempting to obtain federal funding for therapeutic psychedelic studies. One respondent wrote, “I had extensive discussions with NIH institutes about submitting other [grant applications for therapeutic psychedelic studies] but was discouraged from doing so. So, it never got to the point of review.” Another respondent noted that a grant application to NIH for a clinical trial cleared by the US Food and Drug Administration had received a “not recommended for further consideration” decision, so it was not reviewed and given an impact score. One respondent wrote, “NIDA, unlike NIMH, has not recognized the importance of transformative effects of psychedelics for drug dependence.” Another respondent observed, “Not only funding agencies, but reviewers used to be very skeptical and critical of these applications, many of which did not reach council for decisions.” However, this respondent struck an optimistic tone about the current federal funding landscape, stating now “there is hope.”

## Discussion

This appears to be the first survey study of investigators submitting grant applications for therapeutic psychedelic studies to US federal agencies. Findings demonstrate that multiple studies proposing to investigate the therapeutic potential of psychedelics, which were primarily in vitro and animal investigations targeting major depressive disorder and substance use disorders, have been submitted to NIH since 1991-1995. Given that the average funding for an NIH grant in 2022 was $592,617 (National Institutes of Health 2022b), the significant career-spanning federal grant funding obtained by respondents indicates this is a highly skilled group of researchers. That at least three grant applications were submitted prior to 2006 signifies there were efforts by some in this group to conduct therapeutic studies of multiple psychedelics before the psychedelic renaissance began. Looking at the data in totality, they suggest that grant applications for therapeutic applications of psychedelics have risen considerably in recent years and, as discussed below, so has NIH funding for this line of research.

16.7% of these grant applications were funded, which is somewhat lower than the average annual funding rate of 23.4 ± 4.9% for R-01 equivalent grant applications made to NIH from 1998 (the earliest year for which data are publicly available online) to early 2023 (United States National Institutes of Health 2023). Notably, no submitted applications were funded from the 1991-1995 block to the 2001-2005 block and the average annual funding rate for R-01 equivalent grant applications made to NIH from 1998 to 2005 was 29.6 ± 3.3%. Using the assumptions previously discussed in the results section, the funding rate in this sample was somewhere from 19.05% to 22.2% from 2006 to early 2023, which is close to the average annual NIH funding rate for R-01 equivalent grant applications over this period of 20.6 ± 1.9%. This suggests that there has been a considerable increase in NIH support for this line of research since 2006-2010.

Respondents reported important information about the perceived quality of grant applications by NIH reviewers via the best impact scores of funded and unfunded applications. NIH’s website provides the following general guidance about the relationship between impact score and likelihood of receiving funding, “The normalized average of all reviewer impact/priority scores constitutes the final impact/priority score. Impact scores run from 10 to 90, where 10 is best. Generally, impact/priority scores of 10 to 30 are most likely to be funded; scores between 31 and 45 might be funded; scores greater than 46 are rarely funded. Before 2009, NIH used a different score system, with final scores from 100 to 500, where 100 was best (National Library of Medicine 2022).” Of note, respondents reported only impact scores consistent with the post-2009 scoring system. For rejected NIH applications where an impact score was reported, two scores fell within the 10-30 range, one within the 31-45 range, and one was greater than 46. This impact score distribution indicates that despite being considered high quality by reviewers, some applications were not funded by NIH. Notably, one respondent also reported NIH staff had discouraged them from submitting grant applications for therapeutic psychedelic studies and another alleged that reviewers in the past tended to be overly critical of these applications, preventing some from ever reaching committee review.

Respondents predominantly felt grant applications for therapeutic psychedelic studies were significantly less likely to receive federal funding compared to other applications they had submitted, which is notable given that respondents reported career-spanning grant funding consistent with having obtained multiple grants. They also largely believed the US federal government is significantly underinvested in therapeutic psychedelic research. However, most respondents believed the likelihood of obtaining federal funding for therapeutic psychedelic research has improved over the last five years, consistent with this study’s previously mentioned findings.

Some respondents reported that proposed projects not receiving federal funding remained unfunded by any source, though most respondents obtained financial support from elsewhere for these projects, primarily from their own institutions or psychedelic-focused non-profit organizations. While philanthropic funding has been essential to re-launching therapeutic psychedelic research, some researchers have also observed that it is increasingly hard to obtain due to growing interest among potential donors in investing in psychedelic-related commercial endeavors (Powell 2021).

Overall, this study suggests that a tangible shift in NIH support for studies investigating therapeutic applications of psychedelics has occurred in recent years. This is a hopeful finding since, in addition to allowing more investigators to participate in studies on the therapeutic applications of psychedelics, increased public funding for this research could allow for larger clinical trials with longer-term follow-up to better understand the long-term safety profile and durability of response of psychedelic treatments (Hall 2021).

### Limitations

This study is limited by its small sample size. Though any investigator who had submitted a federal grant application for a therapeutic psychedelic study was invited to participate, it is possible that those who had multiple applications rejected by federal agencies may have been more motivated to participate due to frustration by their failed attempts to garner funding. Therefore, the proportion of unsuccessful to successful grant applications may not be an accurate reflection of reality. However, a review of publicly available data on NIH funded grants reveals only a small number of therapeutic psychedelic studies, with very few involving human subjects, though the number has increased in recent years, consistent with this study’s findings.

## Conclusions

This study provides first of its kind data indicating that since at least the early 1990s there have been several grant applications proposing to investigate therapeutic applications of psychedelics submitted to NIH. These data indicate that such grant applications began to rise significantly starting in 2006. While no relevant grant applications submitted prior to 2006-2010 were funded by NIH, the funding rate of applications since that time is close to the average annual NIH funding rate for R-01 equivalent grant applications. Unfortunately, these data do not tell us when the first relevant grant application from researchers in this sample was funded beyond the fact that it was some time after 2005.

## Data Availability

All data produced in the present study are available upon reasonable request to the authors

